# Occupational Exposures and Programmatic Response to COVID-19 Pandemic: An Emergency Medical Services Experience

**DOI:** 10.1101/2020.05.22.20110718

**Authors:** David L. Murphy, Leslie M. Barnard, Christopher J. Drucker, Betty Y. Yang, Jamie M. Emert, Leilani Schwarcz, Catherine R. Counts, Tracie Y. Jacinto, Andrew M. McCoy, Tyler A. Morgan, Jim E. Whitney, Joel V. Bodenman, Jeffrey S. Duchin, Michael R. Sayre, Thomas D. Rea

## Abstract

**Background:** Rigorous assessment of occupational COVID-19 risk and personal protective equipment (PPE) use are not well-described. We evaluated 9-1-1 emergency medical services (EMS) encounters for patients with COVID-19 to assess occupational exposure, programmatic strategies to reduce exposure, and PPE use.

**Methods:** We conducted a retrospective cohort investigation of lab-confirmed COVID-19 patients in King County, WA who received 9-1-1 EMS responses from February 14, 2020 to March 26, 2020. We reviewed dispatch, EMS, and public health surveillance records to evaluate the temporal relationship between exposure and programmatic changes to EMS operations designed to identify high-risk patients, protect the workforce, and conserve PPE.

**Results:** There were 274 EMS encounters for 220 unique COVID-19 patients involving 700 unique EMS providers with 988 EMS person-encounters. Use of “full” PPE including mask, eye protection, gown and gloves (MEGG) was 67%. There were 151 person-exposures among 129 individuals, who required 981 quarantine days. Of the 700 EMS providers, 3 (0.4%) tested positive within 14 days of encounter. Programmatic changes were associated with a temporal reduction in exposures. When stratified at the study encounters midpoint, 94% (142/151) of exposures occurred during the first 137 EMS encounters compared to 6% (9/151) during the second 137 EMS encounters (p<0.01). By the investigation’s final week, EMS deployed MEGG PPE in 34% (3579/10,468) of all EMS person-encounters.

**Conclusion:** Less than 0.5% of EMS providers experienced COVID-19 illness within 14 days of occupational encounter. Programmatic strategies were associated with a reduction in exposures, while achieving a measured use of PPE.

## INTRODUCTION

The first case of 2019 novel coronavirus disease (COVID-19) in King County, Washington was reported on February 28, 2020. Incidence rose exponentially in subsequent weeks.^1^ Emergency medical services (EMS) are the front line of the healthcare system, responding with incomplete information to provide care in heterogeneous, often uncontrolled, circumstances. The COVID-19 pandemic challenges healthcare worker safety in part because of limited supplies of personal protective equipment (PPE). Ideally, EMS strategies would incorporate COVID-19 risk assessment and target use of the limited PPE resource in order to achieve EMS provider safety, extend the supply of PPE, and support high-quality patient care.

The US Centers for Disease Control and Prevention (CDC) established criteria for COVID-19 testing and case management based on history and recent travel to a high-risk area, contact with known or suspected COVID-19 cases, and presence of fever and signs/symptoms of lower respiratory illness.^2^ Based on national guidelines, our regional EMS system initially adopted a screening framework based on travel, exposure to known cases, and specific symptoms. During the initial days and weeks of the outbreak, we identified long-term care facilities (LTCF) as high-risk locales and appreciated the atypical presentations involving COVID-19 illness.^3,4^ As a consequence, we implemented a series of iterative protocol changes with regard to COVID-19 risk assessment and PPE use based on the patient’s clinical profile and response location.

We evaluated all 9-1-1 EMS responses to COVID-19 patients to (1) determine occupational exposure, related workforce quarantine, and potential transmission, and (2) understand how programmatic changes influenced occupational exposure, workforce quarantine, and PPE use amidst the COVID-19 outbreak in Seattle and King County.

## METHODS

### Study Design and Setting

The study is a retrospective cohort investigation of EMS providers responding to 9-1-1 calls for lab-confirmed COVID-19 positive patients in King County, Washington between February 14, 2020 and March 26, 2020. The first US case was documented in neighboring Snohomish County on January 20th, with unrecognized transmission of COVID-19 until clinical diagnosis within King County in late February, 2020.^5,6^ EMS providers who cared for COVID-19 patients were monitored through April 9, 2020 to complete a 14-day surveillance after the final patient encounter date. During this time, COVID-19 disease was defined by the State of Washington as positive reverse transcriptase-polymerase chain reaction (RT-PCR) testing for severe acute respiratory syndrome coronavirus 2 (SARS-CoV-2).

King County is a metropolitan region, covering 2,132 square miles, with 2.2 million persons who reside in urban, suburban, and rural areas. The primary 9-1-1 medical response in King County is two-tiered. The first tier is provided by firefighter emergency medical technicians (EMT). Paramedics comprise the second tier and are dispatched in cases of more severe illness. There are 28 first-tier fire departments and five overarching second-tier paramedic agencies that collectively provide primary emergency response to all 9-1-1 medical calls. In general, stable patients are transported via fire department or private ambulance basic life support units, and more acute patients are transported by advanced life support paramedic units. All EMS, fire, and private ambulance agencies in King County participated in this study. Collectively, there are approximately 4,000 EMS providers in King County. The study was approved by the University of Washington Institutional Review Board.

### Study Population: COVID-19 Encounter, Exposure, and Quarantine/Isolation

The study population are EMS providers who cared for patients with confirmed COVID-19 by RT-PCR tests. EMS is administered by Public Health–Seattle & King County, enabling direct engagement between EMS and Public Health to undertake COVID-19 surveillance. To identify EMS encounters with COVID-19 patients, we linked local and state COVID-19 surveillance systems with EMS electronic records using the patient’s name and date of birth. Patient encounters were included if they occurred within a hierarchical, predetermined transmission window of 3 days prior to symptom onset (if known) or 14 days prior to or after the diagnosis date. Each match was independently verified by an epidemiologist and physician.

A physician reviewed each matched encounter for potential EMS exposure in the electronic health record. If the documented PPE was not a complete ensemble of mask, eye protection, gown, and gloves (MEGG), the case was further investigated by the EMS agency’s appointed health officer (Figure 1). Health officers contacted individuals with possible exposure to understand the specific circumstances of patient involvement and clarify PPE use. The health officer in consultation with physician leadership then made the final determination of exposure and whether quarantine or isolation was indicated according to the CDC risk assessment matrix.^7^

**Figure 1:**
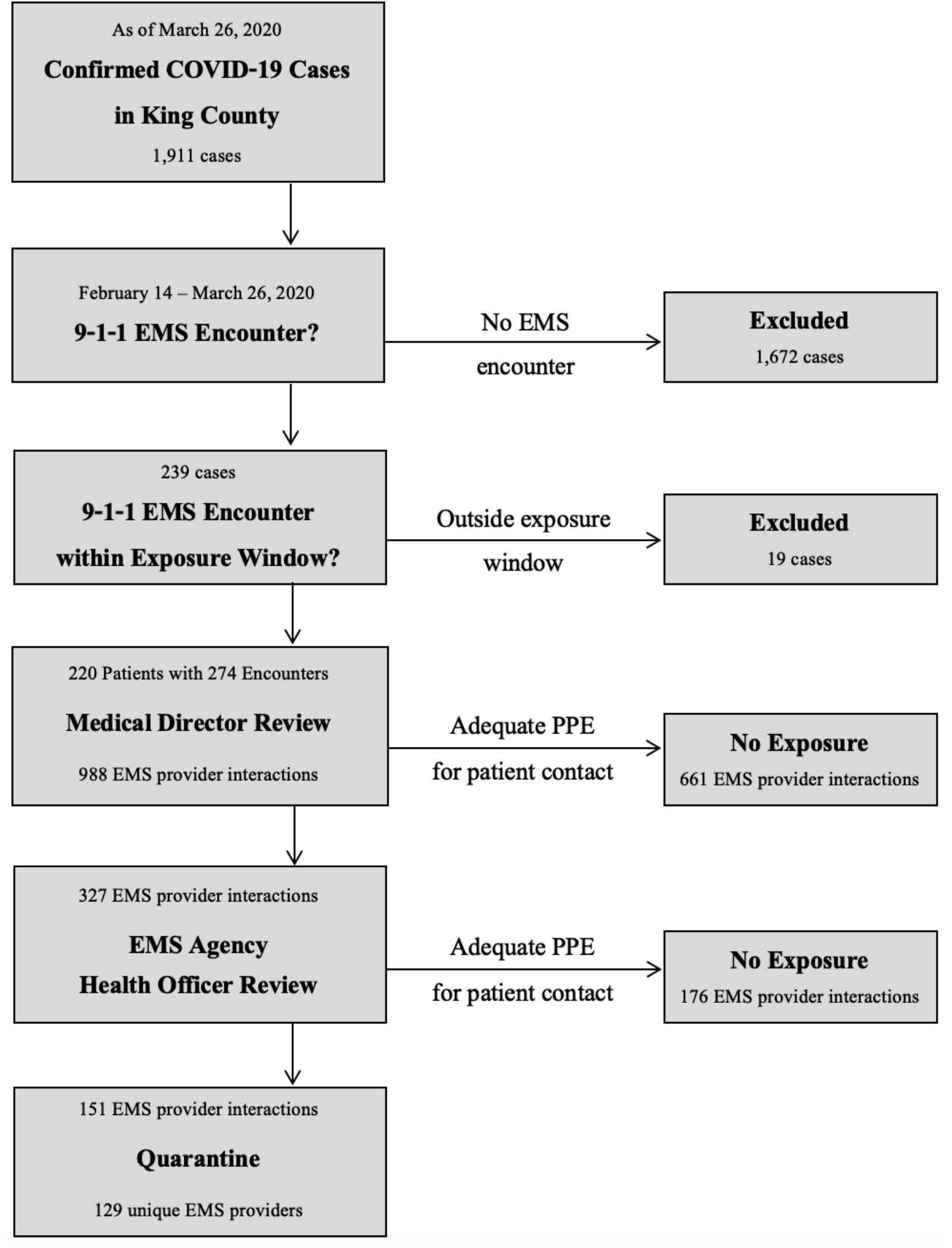
Flow Diagram

An encounter was defined as a 9-1-1 EMS response to a patient confirmed to have COVID-19. An occupational exposure to COVID-19 was defined as a provider-level encounter with inadequate PPE for the patient contact. In addition to eye protection and gloves, a surgical mask was judged to be sufficient for routine patient encounters. However, an N95 mask was required PPE for aerosol generating procedures. For any physical contact with the patient, a gown was required.

EMS agencies implemented regular employee symptom screening upon arrival at work and during the shift. Anyone who felt unwell for any reason returned home until they were asymptomatic and fit for duty per their agency return to work guidelines. EMS providers who became ill regardless of exposure status were deemed symptomatic, placed on isolation, and prioritized for COVID-19 RT-PCR testing through dedicated first responder testing sites. These RT-PCR tests were performed by the University of Washington Virology Laboratory using an assay shown to have a low false negative rate.^8^ Each EMS agency assessed quarantined providers daily. The current investigation used information from both the health officer monitoring program and the Public Health surveillance to ascertain any COVID-19 tests performed among the EMS provider cohort.

### Interventions

#### *(a)* Initial High-Risk Criteria

Prior to the first lab-confirmed case of COVID-19 in King County on February 28, 2020, EMS medical direction issued directives for COVID-19 screening and patient care on February 6 and February 27, 2020. Beginning March 4, EMS providers were advised to don full MEGG PPE if COVID-19 screening included (1) a person with febrile respiratory illness AND travel from an endemic area (initially Wuhan, then broadened to China, South Korea, Iran, or Italy) OR (2) febrile respiratory illness AND known contact with a confirmed COVID-19 patient.

#### *(b)* Dispatch PPE Advised

After February 28, EMS updated the high-risk criteria to include the first LTCF where initial cases were identified, with dispatch to alert “PPE advised” for any response to the address. After additional cases were identified at a second LTCF and a dialysis center, these sites were added as high-risk locations for dispatch. A growing list of LTCFs and congregate living centers soon followed. Beginning March 7, EMS began to treat all LTCFs (skilled nursing facilities, assisted living facilities and adult family homes) as high-risk requiring full MEGG PPE, regardless of clinical illness profile.

#### *(c)* Clinical Criteria Profile

With evidence of community transmission, the requirements for travel history or COVID-19 contact were eliminated as criteria to don MEGG PPE during the first week of March. Medical record review determined that EMS COVID-19 patients did not consistently demonstrate a febrile respiratory illness; criteria were expanded to include any respiratory *or* fever symptoms beginning March 11.

#### *(d)* Scout Program

Case review indicated that initial symptom classification—often derived from dispatch reporting—did not adequately characterize illness and the potential for COVID-19 illness. In response, EMS was using large quantities of PPE to address this uncertainty, though the prevalence of confirmed COVID-19 EMS encounters was estimated to be less than 5%.^1^ Hence, EMS leadership implemented a “scout program” beginning March 14 in which one or two EMS providers donned full MEGG PPE and entered the “hot zone” to perform the initial in-person evaluation while additional crew remained in the “cold zone,” maintaining sight or voice contact, with scout responder(s). The scout evaluation informed the need for remaining EMS crew to don PPE to assist. Conversely, risk assessment was often not feasible in high-acuity, time-sensitive cases. All cardiac arrest cases and cases requiring aerosol-generating therapies required full MEGG PPE with N95 masks.

### Data collection and measurements

We used a uniform methodology to review the narrative and formatted data fields from dispatch and EMS records. Dispatch records were abstracted to characterize 9-1-1 patient concern and pre-arrival notifications. EMS records were abstracted to describe patient characteristics, location, initial vital signs, disposition, clinician impression, and PPE use. PPE use was assessed through review of the EMS report narrative and discrete data fields. Following the first recognized case of COVID-19 in King County, the EMS leadership directed reporting of full PPE use in the electronic record. Beginning March 20, mandatory, item-specific PPE reporting became available through the electronic health record (ESO Solutions, Austin, TX) for all EMS responses. EMS provider quarantine dates and results from COVID-19 testing were recorded.

### Outcomes

We evaluated the number of COVID-19 patient encounters, PPE use, consequent exposures due to inadequate PPE, resulting quarantine, and positive COVID-19 tests among EMS providers.

### Analysis

Descriptive analyses were performed at the EMS encounter and EMS provider levels. EMS encounters were stratified by level of transport while provider level assessments were stratified at the chronologic midpoint of EMS encounters. Due to a subset of providers with multiple patient encounters, we report provider level assessments as both total EMS provider encounters and as unique EMS providers. We used logistic regression to determine if calendar time was associated with a temporal trend in adequate PPE use and EMS provider exposure. To estimate the potential conservation of PPE relative to an indiscriminate MEGG PPE deployment strategy (MEGG for all EMS personnel for all calls), we determined the actual PPE use during the week of March 20-26 among the total number of EMS providers involved on 9-1-1 responses. SAS (version 9.4; SAS Institute) was used to conduct analyses.

## RESULTS

### Characteristics of COVID-19 Patients

There were 220 unique patients with confirmed COVID-19 in Seattle and King County with 9-1-1 EMS encounters in the 14 days prior to, and first 28 days after, the sentinel lab confirmed case in King County. Of these 220 individuals, 54 had two EMS encounters for a total of 274 distinct EMS encounters. Half were female (53%), and the mean age was 74 years. The dispatch complaints were heterogenous; difficulty breathing was the most common complaint, accounting for about 25% (Table 1). The mean initial pulse oximetry reading was 90%. The most common EMS impressions included suspected COVID-19 illness (26%), flu-like symptoms (17%), respiratory distress (17%), and weakness (14%).

**Table 1:** Characteristics of 274 EMS encounters with 220 confirmed COVID-19 patients from February 14 - March 26, 2020.

|  | Total EMS encounters (n=274) | BLS Transport (n=180) | ALS Transport (n=31) | Not Transported (n=63) |
| --- | --- | --- | --- | --- |
| Initial Dispatch Complaint, n (%) of column |  |  |  |  |
| Difficulty Breathing | 68 (25) | 44 (24) | 10 (32) | 14 (22) |
| Sick (unknown) | 68 (25) | 52 (29) | 6 (19) | 10 (16) |
| Infectious Diseases | 51 (19) | 35 (19) | 4 (13) | 12 (19) |
| Trauma | 32 (12) | 17 (9) | 2 (7) | 13 (21) |
| Other* | 55 (20) | 32 (18) | 9 (29) | 14 (22) |
| Dispatched as "PPE Advised", n (%) of column | 196 (72) | 136 (76) | 16 (52) | 44 (70) |
| Origin of 911 response, n (%) of column |  |  |  |  |
| Home/Private Residence | 128 (47) | 76 (42) | 16 (51) | 36 (57) |
| Long term care facility | 118 (43) | 86 (48) | 12 (39) | 20 (32) |
| Outpatient clinic | 20 (7) | 14 (8) | 3 (10) | 3 (5) |
| Public/street | 8 (3) | 4 (2) | 0 (0) | 4 (6) |
| Initial Vital Signs, mean (SD) of column |  |  |  |  |
| Systolic blood pressure, mm Hg (n=238) | 128 (27) | 129 (24) | 130 (37) | 125 (33) |
| Heart rate, beats per minute (n=230) | 91 (23) | 94 (20) | 95 (32) | 82 (26) |
| Respiratory rate, breaths per minute (n=238) | 21 (9) | 21 (8) | 26 (11) | 18 (9) |
| Peripheral oxygen saturation, % (n=197) | 90 (10) | 90 (8) | 82 (18) | 93 (7) |
| EMS Provider Impression, n (%) of column |  |  |  |  |
| COVID** | 72 (26) | 53 (29) | 4 (13) | 15 (24) |
| Flu-like symptoms | 47 (17) | 37 (21) | 4 (13) | 6 (10) |
| Respiratory distress | 46 (17) | 27 (15) | 11 (36) | 8 (13) |
| Weakness | 37 (14) | 25 (14) | 0 (0) | 12 (19) |
| Altered mental status | 16 (6) | 8 (4) | 4 (13) | 4 (6) |
| Injury/Pain | 16 (6) | 13 (7) | 1 (3) | 2 (3) |
| Cardiac | 15 (6) | 6 (3) | 6 (19) | 3 (5) |
| Other*** | 25 (9) | 11 (6) | 1 (3) | 13 (21) |
| Any mention of COVID-19 in EMS record, n (%) of column | 169 (62) | 117 (65) | 17 (55) | 35 (56) |

\*Bleeding/pain, cardiac, and stroke/headache.
\*\*COVID-19 impressions were added to the electronic health record as an option on March 8, 2020.
\*\*\*Vaginal hemorrhage, seizures, obvious death, gastrointestinal hemorrhage, epistaxis, dehydration, urinary tract infection, diabetic hypoglycemia, unspecified convulsions, and skin infection.

### Main results

Among the 274 EMS encounters with COVID-19 patients, there were 429 responding units, involving 700 unique EMS providers with a total of 988 EMS provider encounters (Table 2). Based on initial EMS record review, use of PPE during patient contact was full MEGG (66.9%), basic gloves and eye protection (29.3%), delayed application or partial MEGG (3.1%), or unknown (0.7%), resulting in 327 possible EMS provider exposures. After health officer investigation and physician consultation, 151 EMS provider encounters were determined to have an exposure. As a result, there were 129 unique EMS providers placed on quarantine: 107 after a single exposure and 22 with two exposures. Of the 700 unique EMS providers caring for patients with confirmed COVID-19, 3 (0.4%) tested positive during the 14 days following an encounter (Table 3), yet none of these three had a documented occupational exposure.

**Table 2:** Use of PPE and occupational exposures among EMS provider encounters with confirmed COVID-19 patients from February 14 - March 26, 2020.

|  | Total | *Initial 137 EMS encounters with COVID-19 patients | *Subsequent 137 EMS encounters with COVID-19 patients |
| --- | --- | --- | --- |
| Number of EMS provider encounters, n | 988 | 488 | 500 |
| Any mention of COVID-19 in EMS record, n (%) | 133 (49) | 49 (36) | 84 (61) |
| Personal Protective Equipment, n (% of column total) |  |  |  |
| Full MEGG | 661 (67) | 265 (54) | 396 (79) |
| Partial or Delayed MEGG | 31 (3) | 17 (3) | 14 (3) |
| Basic (gloves/eyes) | 289 (29) | 202 (41) | 87 (17) |
| Missing/unknown* | 7 (1) | 4 (1) | 3 (1) |
| EMS provider encounters with an exposure, n (% of column total) | 151 (15) | 142 (29) | 9 (2) |
| EMS providers exposures per 9-1-1 encounter with an exposure, median (IQR) | 3 (2,3) | 3 (2,3) | 2 (2,3) |
| Interval from EMS provider exposure to patient COVID-19 test result, days, median (IQR) | 4 (2,6) | 5 (2,8) | 3 (1,4) |

\* Initial encounters occurred between February 19 - March 15, 2020 while subsequent encounters occurred between March 16 - March 26, 2020.

**Table 3:** Occupational exposures, quarantine and testing of unique EMS providers with confirmed COVID-19 patient contact from February 14 - March 26, 2020.

|  | Total | *Initial 137 EMS encounters with COVID-19 patients | *Subsequent 137 EMS encounters with COVID-19 patients |
| --- | --- | --- | --- |
| Unique EMS providers** | 700 | 341 | 382 |
| Unique EMS providers with patient exposure(s), n (% of column total) | 129(18) | 121 (35) | 8 (2) |
| Number of exposure(s) for each unique EMS provider, n (% of column total N=700) |  |  |  |
| 0 | 571 (82) | 220 (65) | 374 (98) |
| 1 | 110 (16) | 103 (30) | 7 (2) |
| 2 | 16 (2) | 15 (4) | 1 (<1) |
| ≥3 | 3 (<1) | 3 (1) | 0 (0) |
| Interval from exposure to EMS provider quarantine, days, median (IQR) | 6 (3,9) | 6 (3,9) | 3 (2,3) |
| Total EMS provider quarantine days resulting from exposure(s), n (% of row) | 981 (100) | 951 (97) | 30 (3) |
| COVID-19 testing of unique EMS providers regardless of exposure status, n (% of column) |  |  |  |
| No symptoms reported (Not tested) | 657 (94%) | 312 (91%) | 368 (96%) |
| Symptoms reported | 43 (6%) | 29 (9%) | 14 (4%) |
| Positive | 3 (<1%) | 1 (<1%) | 2 (1%) |
| Negative | 40 (6%) | 28 (8%) | 12 (3%) |

\* Initial encounters occurred between February 19 - March 15, 2020 while subsequent encounters occurred between March 16 - March 26, 2020.
\*\*23 providers were represented in both categories.

The series of practice changes involving dispatch advisement, patient COVID-19 risk criteria, and initial EMS scene deployment were associated with a temporal increase in adequate PPE use and conversely a decrease in EMS provider exposures (Figure 2, p<0.01). When stratified at the encounters midpoint, 94% (142/151) of exposures occurred during the first 137 EMS encounters compared to 6% (9/151) during the second 137 EMS encounters (Table 2, p<0.01).The number of EMS providers quarantined each day increased to a peak of 69 on March 13th and then declined (Figure 3).

**Figure 2:**
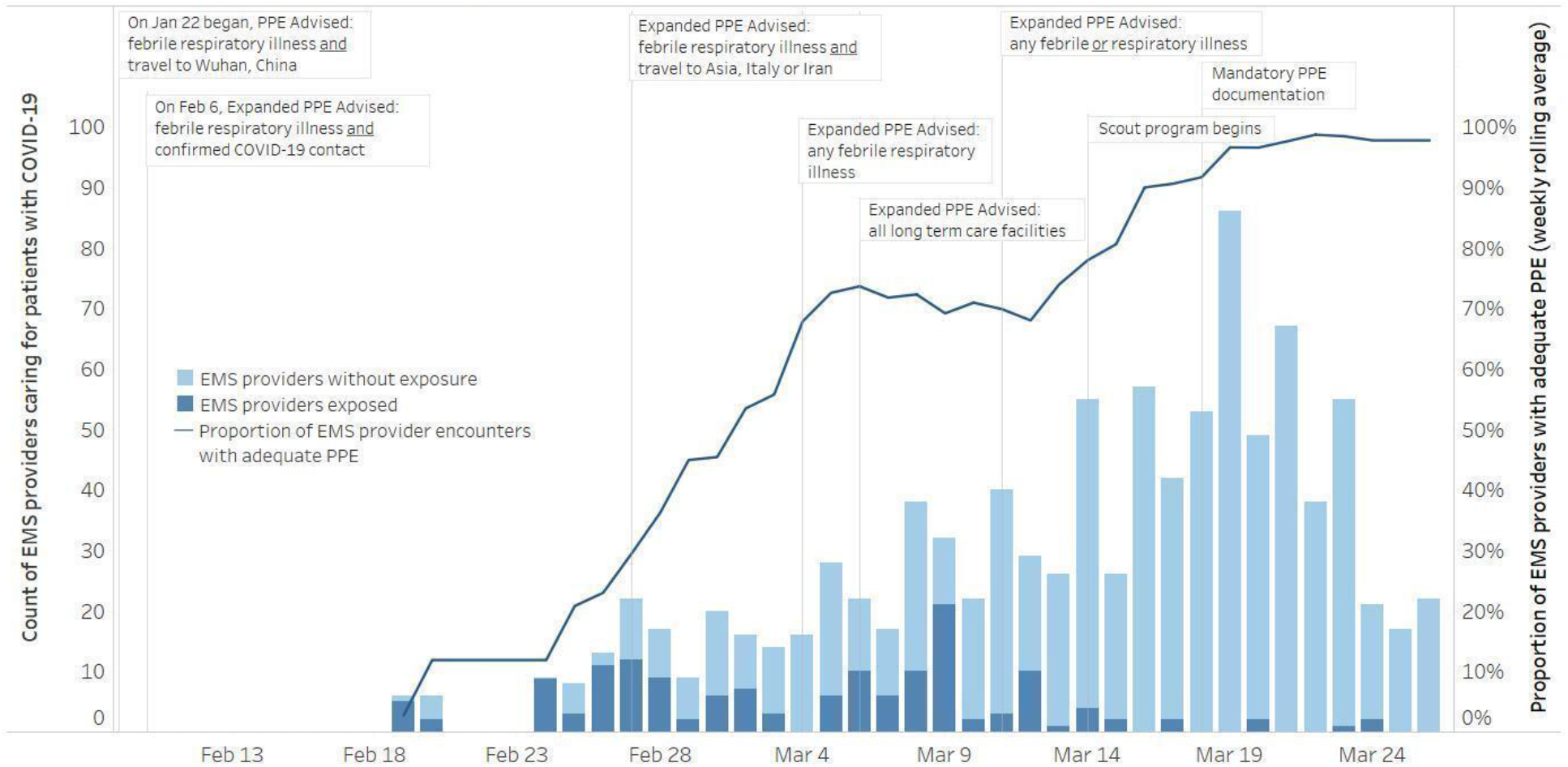
Occupational Exposures and PPE use among EMS Providers caring for COVID-19 Patients, Seattle & King County through March 26, 2020.

**Figure 3:**
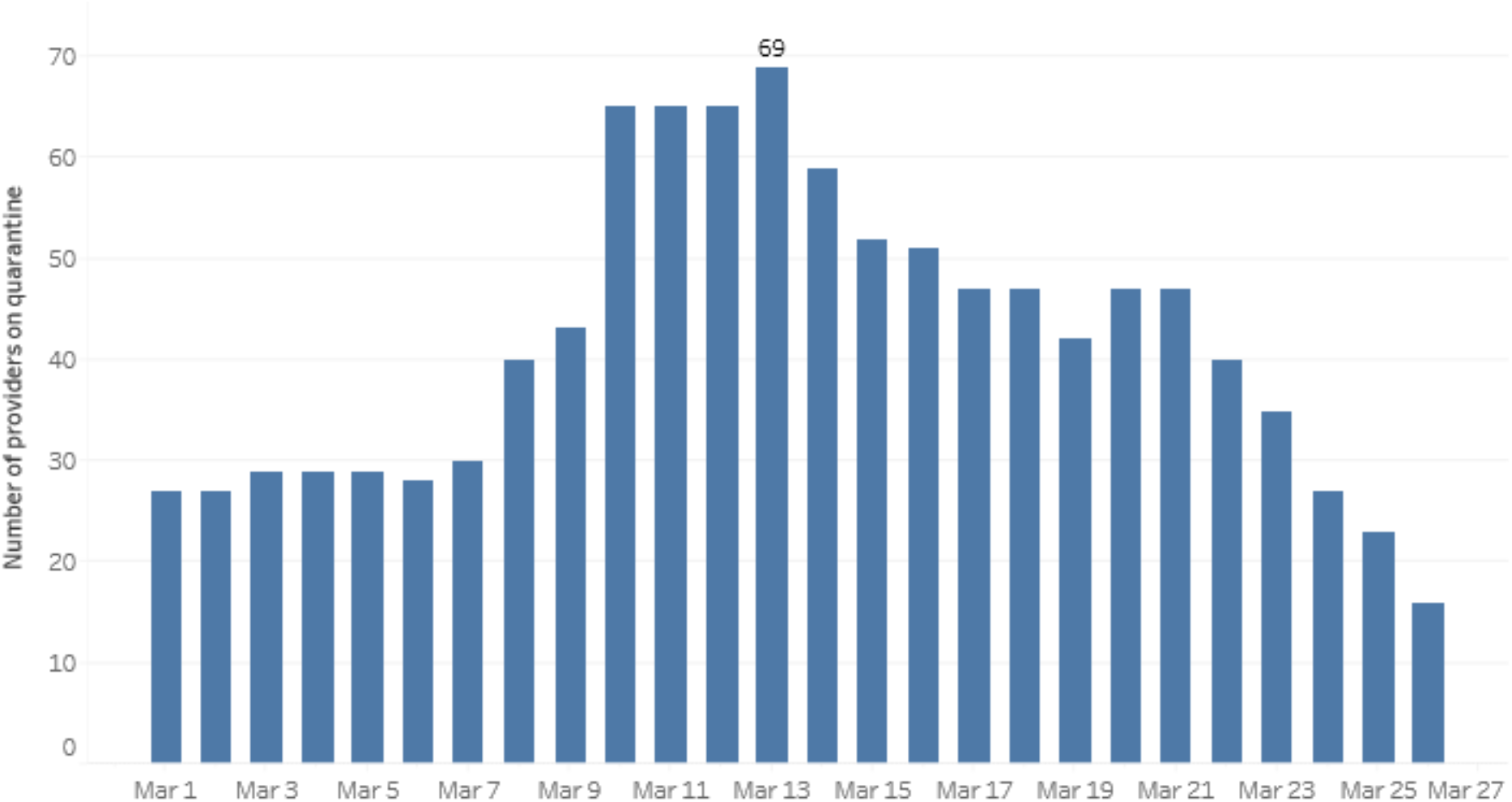
Number of Seattle & King County EMS providers in quarantine by calendar day.

During the final week of the study (March 20-26), there were a total of 3,704 EMS incidents involving 10,468 EMS providers. Of the 10,468 opportunities for PPE deployment, MEGG PPE was used in 3,579 (34%) EMS provider encounters.

## DISCUSSION

In this population-based observational investigation of 274 EMS encounters for patients with COVID-19 involving nearly 1,000 EMS provider-encounters, three EMS providers subsequently tested positive for COVID-19 during the 14 days following the patient encounter. Iterative dispatch and operational EMS responses to COVID-19 risk identification and PPE use were associated with both a temporal decrease in EMS provider COVID-19 exposure and conservation of PPE. Based on these programmatic efforts, full MEGG PPE was deployed in about one-third of all potential EMS provider uses by the end of the study period.

Although healthcare workers (HCW) seem to be at higher risk to contract COVID-19, rigorous assessment of exposure and transmission is largely lacking. Epidemiological reports from China and Italy highlight the substantial burden of illness in HCWs.^9-11^ Locally, in Washington State, a large portion of LTCF staff tested positive for COVID-19.^3^ A preliminary report from CDC regarding the burden of COVID-19 infection among US healthcare personnel suggest HCWs account for 11-19% of national case burden, but did not discern specific type of employment or evaluate the potential source of exposure.^12^ Other reports involving high-risk circumstances involving aerosolizing procedures however have not observed substantial rates of transmission to HCWs.^13^ Similar to our findings, a Taiwanese study reported a secondary attack rate of 0.9% among the subset of COVID-19 exposures occurring in the healthcare setting.^14^ None of these experiences have reported risk to EMS providers, though EMS care appears to be integral for sicker COVID-19 patients. In the 2009 SARS outbreak, the overall incidence of infection was 1.3% in the Taiwanese EMS workforce, which was >100-fold higher than the general public.^15^

In the current investigation, EMS had substantial involvement with COVID-19 illness. The 220 patients represented 14% of all COVID-19 diagnoses in King County, WA through March 26. EMS was typically involved in care for older adults who often presented with heterogeneous symptoms and a range of clinical presentations. COVID-19 in King County was first detected in a clinical population not considered high-risk according to national guidelines at that time, which accounted in part for the fact that 18% of EMS providers in the study had an exposure. Indeed, 85.4% of patients had not been diagnosed with COVID-19 at the time of their EMS encounter.

The high rate of quarantine early on motivated the EMS system to move quickly to adapt to the evolving clinical features and local epidemiology of the COVID-19 outbreak. EMS leadership engaged dispatch and operations to expand COVID-19 risk criteria and to stage patient assessment. The set of measures was associated with a marked reduction in the risk of exposure over the course of investigation. Certainly, there was a learning curve that may have also contributed to reduction in exposure. The collective effect appears to be a temporal reduction in EMS worker quarantine, even though the number of provider encounters with COVID-19 increased over time (Figure 2).

We observed that three of the 700 EMS providers (0.4%) with COVID-19 encounters subsequently tested positive for COVID-19. One case occurred at the outset of the outbreak with onset of provider illness occurring on the same date of COVID-19 encounter. The CDC investigated this case and determined that the 9-1-1 incident that qualified the provider for study inclusion was not responsible for disease transmission. Nonetheless, the provider may have had a patient exposure in the days prior to identification of COVID-19 cases in King County, as further review confirmed care for patients with acute respiratory illness. The providers in all three cases had MEGG PPE during their qualifying encounters. We cannot determine whether transmission occurred during these patient-specific exposures, other occupational activities, or community transmission.

Overall, the cumulative lab-confirmed prevalence in this EMS cohort of 700 unique providers (0.4%) is comparable to the community prevalence (0.2%) during this time frame.^1^ Taken together, these findings suggest that occupational risk can be relatively low and that protective measures can potentially limit disease transmission. The anecdotal experiences in other regions reporting high rates of COVID-19 among EMS providers may be related to the higher prevalence of disease paired with limited availability and use of PPE.

There is an inherent tension between proactive measures to don adequate PPE and conservation efforts due to limited supplies. If PPE were limitless, then indiscriminate use by all providers for every call would help assure EMS provider protection. However, our system had limited supply that was coupled with uncertainty about the severity and duration of the pandemic. Thus, the EMS system strived to target the use of PPE to risk positive patients. The scout strategy for stable patients enabled more deliberate decisions regarding PPE. In contrast, time-critical events such as cardiac arrest required comprehensive EMS PPE given the need for care prior to evaluating COVID-19 risk. The current targeted strategies for MEGG utilization appear to be a viable means to protect EMS providers and conserve PPE.

The retrospective methodology used to assess PPE is imperfect, relying on documentation and case-specific investigation; the two-stage process however enabled detailed provider interviews to assess potential exposure. Provider documentation may introduce bias, although providers were motivated to accurately document PPE. Providers received training and education on best-practices of donning and doffing of PPE, but there was not a dedicated observer to document the quality of the process. The study could not report on the temporal use of PPE across the system, but rather the status after implementation of various interventions designed to better assess COVID-19 risk and responsibly use PPE. Documentation of quarantine evolved during the study period to use a central monitoring database. Thus, quarantine decisions early in the outbreak may be an underestimate of quarantine.

We relied on the statewide Washington Disease Reporting System database to identify COVID-19 positive patients. There likely were patients ill with COVID-19 who interfaced with EMS but were not tested. Alternatively, EMS encounters with COVID-19 positive patients may exist that were not captured due to failed linking of identifiers between EMS and surveillance databases. The study relied on EMS agency health officers and the Washington Disease Reporting System database to identify EMS providers tested for COVID-19. Although unlikely, this dual approach may have missed a lab-confirmed infection in an EMS provider. EMS providers may also have chosen not to get tested or had asymptomatic infection, though symptomatic providers were motivated to be tested and had prioritized access to testing. We cannot confirm the source of the infectious exposure—patient-specific, other occupational, or community transmission—among the few providers with positive tests.

In conclusion, less than 0.5% of EMS providers experienced COVID-19 illness within 14 days of caring for a patient with lab-confirmed COVID-19. Programmatic risk mitigation strategies were associated with a reduction in occupational exposures to COVID-19 among EMS providers, while achieving a measured use of PPE.

## Data Availability

All data relevant to the study are included in the article.

## ACKNOWLEDGEMENTS

We wish to acknowledge Public Health - Seattle & King County, the Washington State Department of Health, the Centers for Disease Control, and the telecommunicators and EMS professionals of Seattle and greater King County.

## REFERENCES

1. Public Health-Seattle & King County. COVID-19 data dashboard. (https://kingcounty.gov/depts/health/communicable-diseases/disease-control/novel-coronavirus/data-dashboard.aspx).

2. Centers for Disease Control and Prevention. Update and interim guidance on outbreak of coronavirus disease 2019 (COVID-19). February 28, 2020 (https://emergency.cdc.gov/han/2020/han00428.asp).

3. Guan WJ, Ni ZY, Hu Y, et al. Clinical Characteristics of Coronavirus Disease 2019 in China. N Engl J Med. 2020; 382: 1708-1720.

4. McMichael TM, Currie DW, Clark S, et al. Epidemiology of Covid-19 in a Long-Term Care Facility in King County, Washington. N Engl J Med. 2020; 382: 2005-11.

5. Bedford T, Greninger A, Roychoudhury P, et al. Cryptic transmission of SARS-CoV-2 in Washington State. April 16, 2020. (https://www.medrxiv.org/content/10.1101/2020.04.02.20051417v2) (preprint).

6. Public Health-Seattle & King County. First death due to novel coronavirus (COVID-19) in a resident of King County. February 29, 2020. (https://kingcounty.gov/depts/health/news/2020/February/29-covid19.aspx)

7. Centers for Disease Control and Prevention. Interim U.S. Guidance for Risk Assessment and Public Health Management of Healthcare Personnel with Potential Exposure in a Healthcare Setting to Patients with Coronavirus Disease (COVID-19). Centers for Disease Control and Prevention. March 7, 2020. (https://www.cdc.gov/coronavirus/2019-ncov/hcp/guidance-risk-assesment-hcp.html).

8. Long DR, Gombar S, Hogan CA, et al. Occurrence and Timing of Subsequent SARS-CoV-2 RT-PCR Positivity Among Initially Negative Patients. May 8, 2020. (https://www.medrxiv.org/content/10.1101/2020.05.03.20089151v1) (preprint).

9. Wang D, Hu B, Hu C, et al. Clinical Characteristics of 138 Hospitalized Patients With 2019 Novel Coronavirus-Infected Pneumonia in Wuhan, China. JAMA. 2020;323(11):1061–1069.

10. Wu Z, Mcgoogan JM. Characteristics of and Important Lessons From the Coronavirus Disease 2019 (COVID-19) Outbreak in China: Summary of a Report of 72 314 Cases From the Chinese Center for Disease Control and Prevention. JAMA. 2020;323(13): 1239–1242.

11. Onder G, Rezza G, Brusaferro S. Case-Fatality Rate and Characteristics of Patients Dying in Relation to COVID-19 in Italy. JAMA. 2020;323(18):1775–1776.

12. Characteristics of Health Care Personnel with COVID-19 -United States, February 12-April 9, 2020. MMWR Morb Mortal Wkly Rep. 2020;69(15):477–481.

13. Ng K, Poon BH, Kiat Puar TH, et al. COVID-19 and the Risk to Health Care Workers: A Case Report. Ann Intern Med. 2020; L20–0175.

14. Cheng HY, Jian SW, Liu DP, et al. Contact Tracing Assessment of COVID-19 Transmission Dynamics in Taiwan and Risk at Different Exposure Periods Before and After Symptom Onset. JAMA Intern Med. 2020 May 1;e202020.

15. Ko PC, Chen WJ, Ma MH, et al. Emergency medical services utilization during an outbreak of severe acute respiratory syndrome (SARS) and the incidence of SARS-associated coronavirus infection among emergency medical technicians. Acad Emerg Med. 2004; 11 (9):903–11.

